# Coronavirus Disease 2019 (COVID-19): Knowledge, attitudes, practices (KAP) and misconceptions in the general population of Katsina State, Nigeria

**DOI:** 10.1101/2020.06.11.20127936

**Authors:** Murtala Bindawa Isah, Mahmud Abdulsalam, Abubakar Bello, Muawiyya Idris Ibrahim, Aminu Usman, Abdullahi Nasir, Bashir Abdulkadir, Ahmed Rufai Usman, Kabir Matazu Ibrahim, Aminu Sani, Ma’awuya Aliu, Shema’u Abba Kabir, Abdullahi Shuaibu, Shafique Sani Nass

**Author notes:** Correspondence to: Dr Murtala B. Isah, Faculty of Natural and Applied Sciences, Umaru Musa Yar’adua University Katsina, PMB 2218, Katsina State, Nigeria.

## Abstract

**Introduction:** Over six million cases of Coronavirus Disease 2019 (COVID-19) were reported globally by the second quarter of 2020. The various forms of interventions and measures adopted to control the disease affected people’s social and behavioural practices.

**Aim:** This study aims to investigate COVID-19 related knowledge, attitudes and practices (KAP) as well as misconceptions in Katsina state, one of the largest epicentres of the COVID-19 outbreak in Nigeria.

**Methods:** The study is a cross-sectional survey of 722 respondents using an electronic questionnaire through the WhatsApp media platform.

**Results:** One thousand five hundred (1500) questionnaires were sent to the general public with a response rate of 48% (i.e. 722 questionnaires completed and returned). Among the respondents, 60% were men, 45% were 25-39 years of age, 56% held bachelor’s degree/equivalent and above and 54% were employed. The study respondents’ correct rate in the knowledge questionnaire was 80% suggesting high knowledge of the disease. A significant correlation (*P* < 0.05) exists between the average knowledge score of the respondents and their level of education (τ_b_ = 0.16). Overall, most of the respondents agreed that the COVID-19 will be successfully controlled (84%) and the Nigerian government would win the fight against the pandemic (71%). Men were more likely than female (*P* < 0.05) to have recently attended a crowded place. Being more educated (bachelor’s degree or equivalent and above vs diploma or equivalent and below) is associated with good COVID-19 related practices. Among the respondents, 83% held at least one misconception related to COVID-19, with the most frequent being that the virus was created in a laboratory (36%). Respondents with a lower level of education received and trust COVID-19 related information from local radio and television stations and respondents at all levels of education selected that they would trust health unit and health care workers for relevant COVID-19 information.

**Conclusion:** Although there is high COVID-19 related knowledge among the sample, misconceptions are widespread among the respondents. These misconceptions have consequences on the short- and long-term control efforts against the disease and hence should be incorporated in targeted campaigns. Health care related personnel should be at the forefront of the campaign.

## Introduction

Coronavirus disease 2019 (COVID-19) caused by the SARS-CoV2 virus has been detected in 213 countries and territories, with over 6 million people infected and over 371 000 deaths as of 31^th^ May 2020. The number of people currently infected in Africa was 144 323 with 4099 deaths. Six out of the top seven countries with most COVID-19 cases and deaths were advanced countries (Worldometer, 2020), thus, the disease is capable of overwhelming even the most advanced healthcare systems. The estimated COVID-19 viral reproduction number (R_0_), a measure of how easily the virus spreads, ranges from 1.4 to 2.5 (WHO, 2020h) or higher (Zhao et al., 2020) which indicates an easily transmittable virus. The virus is transmitted through contact and respiratory routes (WHO, 2020e). The rapid spread of the infection coupled with a short incubation period (2-14 days) (WHO, 2020b) causes an immense burden on the health care system. COVID-19 Case fatality ratios (CFR) of 7.2% in Italy, 2.3 in China and 3.0% in Nigeria have been reported (Onder et al., 2020; WHO, 2020d). The CFR highly fluctuates by country, stage of the pandemic and age group. In Africa, if COVID-19 proceeds unmitigated, an estimated 3.6-5.5 million cases would require hospitalisation among which 52 000 – 107 000 will require intensive care, far more than the burden African healthcare system can handle (WHO, 2020f).

Early intervention in the spread of contagious viral infections is crucial to the control of the disease. In previous epidemics caused by other coronaviruses namely; Severe Acute Respiratory Syndrome Coronavirus (SARS-CoV) in 2003 and Middle Eastern Respiratory Syndrome (MERS), the World Health Organization (WHO) provided recommendations to break the transmission and spread of the diseases. The recommendations include early disease surveillance, case detection and isolation (Alqahtani et al., 2017; Wilder-Smith et al., 2020). Similar recommendations were used for the COVID-19 outbreak. Other recommended guidelines for the general public to prevent COVID-19 spread include voluntary home quarantine, social distancing and other measures such as frequent hand washing and covering of mouth and nose when coughing or sneezing (Ferguson et al., 2020; WHO, 2020a). Currently there is neither a potent vaccine nor recommended medications to treat the disease (WHO, 2020c). As such, it is likely that most of these measures will be sustained to avoid a second wave or resurgence of the disease.

Nigeria recorded its first case of COVID-19 on the 27^th^ February 2020 (NCDC, 2020b) and by the 31^st^ May 2020 the country reported 11 166 confirmed cases and 315 deaths, with the majority of the COVID-19 cases from Lagos state (NCDC, 2020a). Katsina in North-west Nigeria reported 371 cases with 19 deaths in the same period. Since then, the Katsina state government imposed measures such as the closure of all schools, stay at home order and total lockdown of local government areas with an active transmission of COVID-19. While bans on inter-L.G.A. and Inter-state movements were imposed by Katsina state government and the Federal government respectively.

Both the SARS-CoV and MERS epidemics were not reported in Nigeria. However, the emergence of another viral outbreak, Ebola Viral Disease, was accompanied by unusual behaviours and misconceptions (Iliyasu et al., 2015). Similar practices and misconceptions on COVID-19 prompted the WHO to dedicate a domain for myth buster on its website (WHO, 2020c). These include misconceptions such as that the virus is a biological weapon, the virus is not transmitted at higher temperatures and drinking alcohol or hot beverages have a protective benefit against the disease. People’s perception or interpretation of disease outbreaks influences their health care seeking behaviour (Geldsetzer, 2020). As such, COVID-19 control involves understanding the factors associated with people’s behaviour towards the pandemic. For effective control, the uniqueness of various communities needs to be considered, that was why research focusing on diverse communities was required.

North-western Nigeria is the most populated geopolitical zone of the country with a population of 48.9 million, a quarter of the national projected population (National Bureau of Statistics, 2018). The Hausa-Fulani ethnic predominantly inhabits the North-west region of Nigeria and the majority of them are Muslims. Previous outbreaks of diseases such as cholera, poliomyelitis, measles and cerebrospinal meningitis have been recorded in the region (Wakabi, 2008). Notably, unhygienic water and improper hand washing practices were associated with a cholera outbreak in the North-western state of Kano in the mid-90s (Hutin et al., 2003). More than a decade later, unhygienic hand washing practices were the main risk factors associated with a cholera outbreak in villages of the Jigawa state of the region (Gidado et al., 2018). A survey conducted between 2013-2017 found that, up to 73% of people in some rural areas of the region do not practice adequate hand washing practices using soap and water (UNICEF, 2020), and this could have a negative implication for the control of water-borne and respiratory diseases (UNICEF, 2017).

On the other hand, the obstacle for the fight against poliomyelitis in the region was a rejection of the polio vaccine, driven by misconceptions spread amongst the people (Wakabi, 2008). Thus, the frequent lack of trust and resentment towards public health intervention amongst peoples of North-western Nigeria may likely affect the country’s control efforts against COVID-19. Consequently, it became pertinent to assess the level knowledge, attitudes, practices and misconceptions towards the COVID-19 among the people of Katsina State, Nigeria. The outcome of the study will guide further research in the area with the hope to identify key variables to make informed decisions in the fight against the outbreak of COVID-19 by the relevant authorities.

## Materials and Methods

### Study design

Due to the restrictions on movement and lockdown measures imposed on some Local Government Area (LGAs) of the state, the data for the study was collected online. The respondents in this cross-sectional survey were internet users residing in the 34 LGAs of Katsina state. The survey was conducted from 7^th^ May, 2020 to 18^th^ May, 2020. The lockdown order was eased in the state on 19^th^ May 2020, as such data collection for the study was terminated.

### Questionnaire preparation

The questionnaire was divided into sections assessing the demographic characteristics of the sample, knowledge, attitude, practices, misconceptions and source of information related to COVID-19. The questionnaire for the knowledge, attitudes and practices towards COVID-19 was a modification of the questionnaire developed by Zhong et al. (2020), adopted with the Authors’ consent. The questionnaire consisted of 12 items on knowledge (Q1-Q12), two questions on attitude (Q13 and Q14) and three questions on practices (Q15-Q17) (Table 1). The knowledge assessed includes knowledge on clinical presentation (Q1-Q4), transmission routes (Q5-Q7) and prevention and control (Q8-Q12). Three questions on COVID-19 related misconceptions were developed concerning widespread misconceptions circulating among the people since the beginning of the pandemic. Two items on the source of information were adapted from the UNICEF’s Risk Communication and Community Engagement (RCCE) action plan guidance on COVID-19 preparedness and response (WHO, 2020g). A Hausa language translation of the questionnaire (Supplementary materials) was produced. The data collected remained anonymous. The study was approved by the research ethics committee of the State Ministry of Health, Katsina State (REF: MOH/ADM/SUB/1152/1/375).

**Table 1.** Summary of the overall responses to the questionnaire used to assess knowledge, attitude and practices (n = 722)

| Questions | Response |
| --- | --- |
| **Knowledge (options: true, false or I don't know)** |  |
| Q1. The main clinical symptoms of COVID-19 are fever, fatigue and dry cough | 91% correct |
| Q2. Unlike the common cold, stuffy nose, runny nose, and sneezing are less common in persons infected with the COVID-19 virus | 61% correct |
| Q3. There currently is no effective cure for COVID-19, but early symptomatic and supportive treatment can help most patients recover from the infection | 90% correct |
| Q4. Not all persons with COVID-19 will develop to severe cases. Only those who are elderly, have chronic illnesses, and are obese are more likely to be severe cases | 82% correct |
| Q5. Eating or contacting wild animals would result in the infection by the COVID-19 virus | 42% correct |
| Q6. Persons with COVID-19 cannot infect the virus to others when a fever is not present | 62% correct |
| Q7. The COVID-19 virus spreads via respiratory droplets of infected individuals | 87% correct |
| Q8. Ordinary residents can wear general medical masks to prevent the infection by the COVID-19 virus | 86% correct |
| Q9. It is not necessary for children and young adults to take measures to prevent the infection by the COVID-19 virus | 71% correct |
| Q10. To prevent the infection by COVID-19, individuals should avoid going to crowded places such as markets and avoid taking public transportations | 93% correct |
| Q11. Isolation and treatment of people who are infected with the COVID-19 virus are effective ways to reduce the spread of the virus | 96% correct |
| Q12. People who have contact with someone infected with the COVID-19 virus should be immediately isolated in a proper place. In general, the observation period is 14 days | 95% correct |
| **Attitude** |  |
| Q13. Do you agree that COVID-19 will finally be successfully controlled? (options (agree, disagree or I don't know) | 85% agree, 5% disagree, 10% I don't know |
| Q14. Do you have confidence that Nigeria can win the battle against the COVID-19 virus? (options: yes or no) | 71% yes, 29% no |
| **Practices (options: yes or no)** |  |
| Q15. In the last two weeks, have you gone to any crowded place? | 40% yes, 60% no |
| Q16. In recent days, have you worn a mask when leaving home? | 65% yes, 35% no |
| Q17. In recent days, have you washed your hands with soap and water or hand sanitizer regularly? | 89% yes, 11% no |

### Sampling procedure

A non-probability convenience sample of the general population was reached through volunteers recruited from our network of contacts. The volunteers were trained, and they assisted with administering the questionnaire and receiving the responses. An invitation to participate poster was created and shared among the contacts of the data collectors and WhatsApp groups consisting mainly of people residing in Katsina state.

### Data collection

The electronic questionnaires for the study were administered through the WhatsApp social media platform. An introductory message and informed consent statement were sent to the respondents and the questionnaire, transcribed into strings of WhatsApp messages, was forwarded only to those who wished to participate in the survey. Out of the 1500 people contacted, 722 answered the questionnaire and 778 declined or did not respond (response rate = 48%). The responses received were carefully collated using ODK Collect android application. An excel summary of the data was downloaded from the server (https://odk.ona.io/) on the final day of the data collection.

### Data analysis

Statistical analysis was conducted using SPSS V26. Descriptive statistics such as frequencies, percentages, means and standard deviation were used to summarise qualitative data. The association between respondents characteristics and knowledge, attitudes, practices and misconceptions towards COVID-19 were analysed using chi-square test or Fisher’s exact test for categorical variables. While, the Mann-Whitney U test or Kruskal Wallis test followed by Dunn’s multiple comparison test were used to analyse discrete non-parametric data. The Kendall’s τ coefficient was used to estimate correlation between variables. Multiple logistic regression was used to determine the factors significantly associated with practices among the demographic variables. The best model was constructed using a backward stepwise approach. Statistical significance was considered at *P* value < 0.05.

## Results

Table 2 summarises the demographic characteristics of the sample. Out of the total number of the respondents, 59.6% were male, 55.1% were married and mainly residents of the Katsina senatorial zone. The respondents were predominantly educated (with 56% having a bachelor’s degree/HND or above), < 40 years of age and were mainly students (29.9%), employed in service (29.9%) or engaged in physical labour, self-employment or local business (24.4%).

**Table 2.** Demographic characteristics of the sample (n= 722)

| Variable (n) | Summary value (% total) |
| --- | --- |
| **Gender** |  |
| Male (430) | 59.6 |
| Female (292) | 40.4 |
| **Age group** |  |
| 16-24 (269) | 37.3 |
| 25-39 (327) | 45.3 |
| 40 and above (126) | 17.5 |
| **Marital status** |  |
| Single (398) | 55.1 |
| Ever married (324) | 44.9 |
| **Education level** |  |
| None/others (22) | 3.0 |
| Junior secondary school and below (35) | 4.8 |
| Senior secondary school (110) | 15.2 |
| Diploma/NCE (151) | 20.9 |
| HND/Bachelors degree (308) | 42.7 |
| Master's/Professional degrees and above (96) | 13.3 |
| **Occupation** |  |
| Physical labour/self-employed/local business (175) | 24.5 |
| civil servant level 12 and below (155) | 21.5 |
| civil servant level 13 and above/retired (61) | 8.4 |
| Student (216) | 29.9 |
| Housewife (53) | 7.3 |
| Unemployed (62) | 8.6 |
| **Place of residence** |  |
| Daura senatorial zone (83) | 11.5 |
| Funtua senatorial zone (163) | 22.6 |
| Katsina senatorial zone (475) | 65.8 |

### COVID-19 knowledge

The respondents answered nine out of the 12 questions (Q1-Q12) on COVID-19 knowledge with a correct rate of >70% (Table 1). The three questions with the least correct rate were Q5 and Q6 which assessed COVID-19 related knowledge on mode of transmission and Q2 which assessed knowledge of clinical symptoms. The average correct rate (all questions) for the sample was 80% (SD 16%, range 0%-100%). Among the independent variables recorded, level of education significantly correlated with average knowledge scores (τ_b_ = 0.16, *P* < 0.05).

### COVID-19 related attitudes

We asked two questions to assess the attitude of the respondents on the final success in COVI-19 control and the ability of the Nigerian government to fight the pandemic. Overall, the majority of the respondents agreed that the COVID-19 would be successfully controlled (84%) and the Nigerian government will win the fight against the pandemic (71%). However, across all demographic variables, a higher proportion of the respondents agreed to final success in control than to the government’s ability to battle the pandemic (Table 3). The latter attitude significantly differed with the age of the respondents (*P* < 0.001). The respondents in the 25-29% age group have the least confidence in the government (34.25% no confidence) compared to other age groups.

**Table 3.**
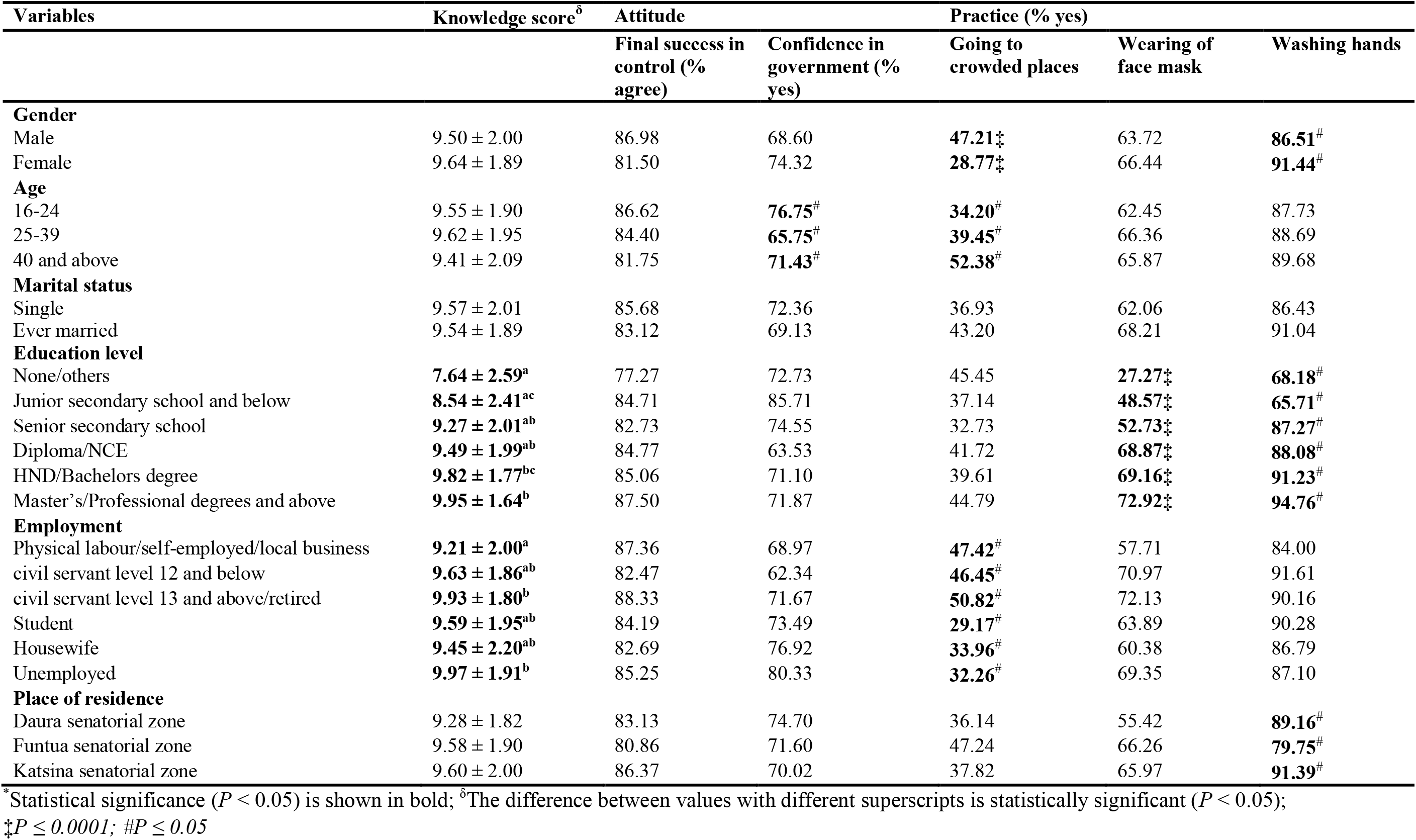
Knowledge, attitude and practices towards COVID-19 among the study respondents (n = 722)^*^

### COVID-19 related practices

About 47% of men in the sample admitted to attending crowded places within the last two weeks, significantly more than women (29% visited crowded places) (*P* < 0.0001) (Table 3). Respondents in the age group of above 40 years were more likely to have attended a crowded place compared to other age groups (*P* < 0.05). Furthermore, employment status significantly affected the response to the question (*P* < 0.05). Civil servants above level 13 (51%), civil servants below level 12 (47%) and respondents in physical labour/self-employment/local business (47%) attended crowded places more than the respondents in the other employment categories. Multiple binary logistics analysis revealed that the male gender was significantly associated with visiting crowded places (vs female, OR = 2.25, *P* < 0.0001) (Table 4).

**Table 4.** Multiple binary logistic regression analysis of factors significantly associated with COVID-19 related practices

| Variable | OR (95% CI) | P value |
| --- | --- | --- |
| **Going to crowded places (yes)** |  |  |
| Gender (male vs female) | 2.25 (1.53 – 3.23) | <0.0001 |
| **Washing of hands regularly (yes)** |  |  |
| Education (Bachelor's degree/HND vs Diploma/NCE or lower) | 2.11 (1.28-3.56) | 0.0042 |
| Education (master's and above vs Diploma/NCE or lower) | 4.40 (1.81-13.26) | 0.0030 |
| Place of residence (Funtua vs Katsina senatorial zones) | 0.36 (0.22-0.61) | 0.0001 |

The level of education significantly affected whether the respondents answered with no on Q16 (wearing a mask while leaving home) and Q17 (washing hands regularly). Only 28% of the respondents in the none/other level of education reported wearing a mask when leaving home. Respondents with a master’s degree or higher level of education have the highest rate of face mask use and washing of hands regularly. Furthermore, the gender and place of residence of the respondents significantly affected the practice of regular hand washing (*P* < 0.05). In a multiple logistic regression analysis, having Bachelor’s degree/HND or higher level of education was significantly associated with an increased rate of hand washing (Bachelor’s degree/HND vs Diploma/NCE or lower, OR = 2.11; Master’s degree and above vs Diploma/NCE or lower OR = 4.4, *P < 0*.*05*), while living in Funtua zone is associated with the reduced rate (vs Katsina senatorial zone, OR = 0.36, *P* = 0.0001).

### COVID-19 related misconceptions

Out of the 722 study respondents, only 122 (17%) answered all three questions on misconceptions with a definitive “no” (Table 5). Thus, 83% of the respondents held at least one or did not know the truth about a COVID-19 misconception. Among the misconceptions, 36% of the respondents believed that COVID-19 was created in a laboratory and 33% thought it was to depopulate the world. Having no misconception was significantly affected only by the level of education (*P* < 0.0001) (Table 6). The misconception towards COVID-19 pandemic was highest in respondents in the none/other category of the level of education (95%) and lowest in respondents with master’s and above (76%). The average COVID-19 knowledge was 9.76 ± 1.76 in the respondents with no misconception and 9.52 ± 1.99 in the respondents who held COVID-19 related misconceptions.

**Table 5.** Overall COVID-19 related misconceptions in the sample (n =722)

| Questions | Yes |  | No<sup>*</sup> |  | I don't know |  |
| --- | --- | --- | --- | --- | --- | --- |
|  | n | % total | n | % total | n | % total |
| Q18. Do you think that COVID-19 was created in the laboratory by a government or terrorist organization? | 257 | 36 | 202 | 28 | 263 | 36 |
| Q19. Do you think that COVID-19 was a result of technological advancement such as 5G? | 135 | 19 | 300 | 42 | 287 | 40 |
| Q20. DO you think that COVID-19 was created to depopulate the world? | 235 | 33 | 220 | 30 | 267 | 37 |

\*Number of respondents who answered all 3 questions with “no” = 122 (17%)

**Table 6.** COVID-19 related misconceptions in the sample by demographic variables and knowledge score

| Variable | Respondents with no misconception |  | Respondents with at least one misconception |  |
| --- | --- | --- | --- | --- |
|  | n | % total | n | % total |
| **Gender** |  |  |  |  |
| Male | 78 | 18.14 | 352 | 81.86 |
| Female | 44 | 15.07 | 248 | 84.93 |
| **Age group** |  |  |  |  |
| 16-24 | 39 | 14.50 | 230 | 85.50 |
| 25-39 | 63 | 19.27 | 264 | 80.73 |
| 40 and above | 20 | 15.87 | 106 | 84.13 |
| **Marital status** |  |  |  |  |
| Single | 66 | 16.58 | 332 | 83.42 |
| Married | 56 | 17.28 | 268 | 82.72 |
| **Education level** |  |  |  |  |
| None/others | 1 | **4.55<sup>‡</sup>** | 21 | 95.45 |
| Junior secondary school and below | 2 | **5.71<sup>‡</sup>** | 33 | 94.29 |
| Senior secondary school | 22 | **20.00<sup>‡</sup>** | 88 | 80.00 |
| Diploma/National certificate of Education | 2 | **13.25<sup>‡</sup>** | 131 | 86.75 |
| HND/Bachelors degree | 54 | **17.53<sup>‡</sup>** | 254 | 82.47 |
| Master's/Professional degrees and above | 23 | **23.96<sup>‡</sup>** | 73 | 76.04 |
| **Occupation** |  |  |  |  |
| Physical labour/self employed/local business | 24 | 13.71 | 151 | 86.29 |
| civil servant level 12 and below | 31 | 20.00 | 124 | 80.00 |
| civil servant level 13 and above/retired | 14 | 22.95 | 47 | 77.05 |
| Student | 37 | 17.13 | 179 | 82.87 |
| Housewife | 06 | 11.32 | 47 | 88.68 |
| Unemployed | 10 | 16.13 | 52 | 83.87 |
| **Place of residence** |  |  |  |  |
| Daura senatorial zone | 14 | 16.87 | 69 | 83.13 |
| Funtua senatorial zone | 23 | 14.11 | 140 | 85.89 |
| Katsina senatorial zone | 85 | 17.86 | 391 | 82.14 |
| **Knowledge score** | 9.76 ± 1.76 |  | 9.52 ± 1.99 |  |

<sup>‡</sup> $P \leq 0.0001$

### Sources of information

The top 10 sources of COVID-19 related information were WhatsApp>other social media>other national TV stations>internet browsing>FM radio stations>international TV stations>local TV stations>health unit/health care worker>family members>friends (Table 7). On the other hand, the top 10 most trusted media source for COVID-19 related information were International TV stations>Health unit/Health care worker>other TV stations>internet browsing>FM radio stations>local TV stations>WhatsApp>Other social media>Other radio stations>family members. The number of channels/sources per respondents in terms of where they receive information was 3.6, whereas the number of trusted channels/sources per respondent was 2.4 (Table 8). WhatsApp and other social media channels were 20% less selected as a trusted source compared to as a source of information on COVID-19. Other channels/sources, except Health unit/Health care worker, were also generally less selected as trusted sources than as sources of COVID-19 related information. Whereas 24% of the respondents received information relating to COVID-19 from Health unit/Health care worker, 27% selected that they trust them as a source. Overall, Health unit/Health care workers were the eighth most frequently reported source of COVID-19 related information by the study respondents but first as a trusted source.

**Table 7:** Sources of COVID-19 related information among the sample

|  | **Q21. What are the major channels or sources do you receive your information on COVID-19 (%)** | **Q22. Which channel/who do you trust the most to receive information related to coronavirus (%)** | **Difference** |
| --- | --- | --- | --- |
| WhatsApp | 36 | 16 | -20 |
| Social Media (not WhatsApp) | 35 | 15 | -20 |
| Other TV stations (National, Channels TV, TVC etc) | 33 | 26 | -7 |
| Internet browsing | 33 | 22 | -11 |
| FM Radio stations | 31 | 21 | -10 |
| International TV stations | 30 | 28 | -2 |
| Local TV stations (KTTV or NTA) | 26 | 18 | -8 |
| Health unit/Health care worker | 24 | 27 | 3 |
| Family members | 23 | 11 | -12 |
| Friends | 23 | 9 | -14 |
| Other radio stations | 19 | 12 | -7 |
| Religious leaders | 13 | 9 | -4 |
| Community health workers | 11 | 10 | -1 |
| Community leaders | 7 | 3 | -4 |
| Any person from the community | 7 | 3 | -4 |
| Other community mobilisers | 5 | 3 | -2 |
| Traditional healers | 2 | 2 | 0 |
| Traditional midwives | 2 | 1 | -1 |

**Table 8.** Sources of COVID-19 related information by education level

|  | Source of information |  | Source of information likely to trust |  |
| --- | --- | --- | --- | --- |
|  | /respondent | Main | /respondent | Main |
| Others/none | 2.7 | • FM radio stations (55%)<br>• Family members (50%)<br>• Local TV stations (27%)<br>• Any person from the community (23%) | 1.8 | • FM radio stations (45%)<br>• Family members/other radio stations (23%)<br>• Local TV stations (18%)<br>• Health unit/Health care Any person from the community (14%) |
| Junior secondary and below | 3.1 | • FM radio stations (40%)<br>• Friends (37%)<br>• Health unit/Health care worker (29%)<br>• Social media/internet browsing (26%) | 2.6 | • FM radio stations (43%)<br>• Health unit/Health care worker (34%)<br>• Social media (23%)<br>• Other radio stations/family (20%) |
| Senior secondary school | 3.3 | • FM radio stations (32%)<br>• WhatsApp/internet browsing (30%)<br>• Friends (29%)<br>• Family (27%) | 2.0 | • International TV stations (24%)<br>• Other TV stations (19%)<br>• Health unit/Health care worker/FM radio stations/local TV stations/internet browsing (16%) |
| Diploma/NCE | 3.4 | • WhatsApp (38%)<br>• FM radio stations/social media (32%)<br>• Local TV stations/other TV stations (27%) | 2.3 | • FM radio stations (25%)<br>• Local TV stations/international TV stations (23%)<br>• Health unit/Health care worker (22%)<br>• Other TV stations/WhatsApp (21%) |
| Bachelor's degree/HND | 3.5 | • Social media (not WhatsApp) (41%)<br>• Internet browsing/WhatsApp/other TV stations (36%)<br>• International TV stations (30%)<br>• Local TV stations (26%) | 2.4 | • Other TV stations (30%)<br>• Health unit/Health care worker/international TV stations (28%)<br>• Internet browsing (27%)<br>• WhatsApp (17%) |
| Master's and above | 5.1 | • International TV stations/WhatsApp (49%)<br>• Internet browsing (48%)<br>• Other TV stations (47%)<br>• Local TV stations (39%) | 3.1 | • International TV stations (46%)<br>• Health unit/Health care (42%)<br>• Other TV stations (36%)<br>• Internet browsing (31%) |
| **Average** | 3.6 | - | 2.4 | - |

FM radio stations were among the main source of information on COVID-19 in the respondents with level of education of Diploma/NCE and below (Table 8). Furthermore, friends or family members were frequently selected as sources of information by the respondents with level of education of senior secondary school or below. On the other hand, respondents with education level of bachelor’s degree/HND and above frequently selected WhatsApp and other social media, internet browsing and TV stations (local, national and international) as sources of information. While the respondents with the level of education of none/others trusted mainly the sources they receive information on COVID-19 from, respondents with a higher level of education trust sources different from where they mainly receive information. Notably, respondents in all levels of education frequently reported that they trust Health unit/Health care workers. Despite this, only respondents in the junior secondary school group have Health unit/Health care workers in the top four sources of COVID-19 related information.

## Discussion

This study assessed the level of COVID-19 related knowledge, attitudes, practices as well as misconceptions in a predominantly Muslim-Hausa society of Katsina state in Nigeria. An overall COVID-19 related knowledge of 80% in the study sample indicated high knowledge of the clinical symptoms, mode of transmission and control measures against the disease. This finding was not surprising since the study was conducted when active COVID-19 control measures, such as the lockdown that directly affected every individual in the state, were active. Similar studies in Nigeria, and countries within Africa (e.g. Tanzania and Ghana (Dorcas et al., 2020; Rugarabamu et al., 2020)) and beyond (e.g. China and Malaysia (Azlan et al., 2020; Zhong et al., 2020)) revealed good COVID-19 knowledge in the study samples that consisted of people with internet access. This finding could reflect the extensive media coverage of the disease and governments’ responses throughout the pandemic. However, one cause for concern on the outcome of the study is the low level of knowledge on COVID-19 transmission by asymptomatic infected individuals (knowledge Q6, 62% correct rate) in the respondents. There have been reports of COVID-19 transmission by asymptomatic (Bai et al., 2020; Yu and Yang, 2020) as well as pre-symtomatic (Arons et al., 2020) individuals, with a negative implication on the fight against the disease (Gandhi et al., 2020). Since the positive response rate to the question (Q6) increased with increasing level of education (Supplementary Table 1) like the average knowledge score (Table 3), enlightenment programmes on asymptomatic transmission should be included in local health promotional campaigns, targeting especially those with lower levels of education.

Okoro et al. (2020) observed that inmates of a custodian centre in Enugu state, Nigeria, that attained higher level of education were more knowledgeable about COVID-19 than other inmates. The positive correlation between the scores of COVID-19 knowledge and education level of the respondents perhaps reflects their source of information on the disease and how they understood the information. In our study, the respondents with the highest average knowledge score, i.e. master’s and above, received COVID-19 related information from diverse channels/sources (an average of 5.1 sources/individual) (Table 8). This finding could mean that their chances of encountering information on the questions asked in the questionnaire are also higher. It may not be feasible to increase the number of channels/sources of information for the respondents with junior secondary school education or lower as a strategy to improve their understanding of the disease. Rather, more information on COVID-19 may be incorporated into the major channels these individuals receive information. These channels are, FM radio stations and local TV stations (Table 8).

Positive attitude towards the final successful control of COVID-19 is high in this study. This optimistic attitude is ubiquitous globally, as shown in studies with Ethiopian (Aynalem et al., 2020), Tanzanian (Rugarabamu et al., 2020), Chinese (Zhong et al., 2020) and Malay (Azlan et al., 2020) populations as well as a multinational study with respondents from six continents (Ali et al., 2020). Thus, a positive attitude on final success is likely unrelated to any demographic characteristic. In studies that used the same questions on attitude with our study, 84% Ethiopian and Paraguayans, 96% Malays and 97% Chinese respondents have confidence that their country will win the battle against COVID-19. In our sample, the rate was 71%. The researchers from China and Malaysia attributed the high level of confidence on the government to the drastic efforts taken by the authorities during previous and current pandemics (Azlan et al., 2020; Zhong et al., 2020). It remains to be investigated whether the individuals in our sample perceived the government’s COVID-19 control efforts negatively. which led to the overall lower confidence in the government. Significantly, we found that bad COVID-19 practices in terms of wearing of face mask and washing of hand were higher in the respondents with a negative attitude towards the government (Supplementary Table 2). Thus, it is critical to address the factors associated with negative attitudes towards the government as this may undermine the fight against COVID-19. Although the respondents with a negative attitude towards the Nigerian government’s ability to control COVID-19 had a lower average knowledge score than those who had confidence (Supplementary Table 3), we do not know whether there is a causal relationship. Thus, we cannot at this stage recommend improving COVID-19 knowledge as a measure to increase confidence in government or vice versa.

Effective COVID-19 control measures rely heavily on good COVID-19 related practices. Social distancing has been recognised as a very important of such practices (Lewnard and Lo, 2020). In this study, men were more likely to have attended a crowded place in the last two weeks, a time when lockdown order was in effect. The majority of these men were above 40 years of age and worked as civil servants or physical labour/self-employed/local business owners. Although we did not ask the respondents on the nature of the crowded place they have attended, these crowds may be unrelated to workplaces, since most businesses were closed and civil servants below level 12 were ordered to work from home. This finding could indicate that older adults were attending social, religious or other forms of gatherings during the lockdown. Thus, considering that this age group is more vulnerable to the fatal effects of COVID-19 (Onder et al., 2020), they should be targeted for enlightenment on proper social and physical distancing etiquettes. The outcome is particularly important for our respondents belonging to the physical labour/self-employed/local business employment group who also have the least frequency of face mask use and frequent hand washing compared to all employment groups (Table 3).

Since the early stages of the pandemic, a lgreat scourge of conspiracy theories appeared online, leading to misinformation and disinformation on the virus (Van Bavel et al., 2020). Misconceptions about the disease may undermine efforts for immediate as well as long-term control measures, especially vaccines (Jolley and Douglas, 2014). In an earlier cross-sectional survey of internet users, 47% of Nigerians held the misconception that COVID-19 was a biological weapon (Olapegba et al., 2020). A larger proportion of our study respondents (83%) held at least one misconception on COVID-19. Respondents with a lower level of education had the highest rates of misconception (Table 6), negative attitudes and bad COVID-19 related practices (Supplementary Table 4) as well as the lowest source of COVID-19 related information (Table 8). Furthermore, they appear to trust any source they receive COVID-19 related information from (Table 8). It is therefore important for local stations to include campaigns that debunk misinformation and disinformation about the disease. This finding is very critical for the future of Nigeria’s fight against COVID-19, considering the previous scepticisms of the North-West Nigeria region regarding vaccines, incidentally, also triggered by similar misconceptions (Ghinai et al., 2013).

## Conclusion

To the best of our knowledge, this is the first study to investigate the KAP and misconceptions towards COVID-19 focused on the Hausa ethnic group. The findings of this study suggest that the study respondents had good knowledge of COVID-19. The effect of the level of education of the respondents on their COVID-19 related practices and the misconceptions they held calls for more research on the general public, which inevitably has a higher proportion of less educated individuals than our sample. We have also identified the main channels the respondents receive COVID-19 related information and the sources they are likely to trust the most, which could be used to deliver targeted information.

### Recommendation

It is recommended that other community engagement strategies, beyond social mobilisation, would be incorporated in the control measures against COVID-19. Factors that need to be considered when communicating and imposing policies should include the current security situations, economic conditions, norms, values and past experience of the communities. Consultations channels aimed at getting feedback from the people, partnership with members of the community and ensuring culturally and religiously appropriate messages may improve cooperation by the people. Measures to increase the peoples’ contact with healthcare units and health care workers on matters associated with COVID-19 are encouraged, considering that majority of the respondents placed trust on them for information. Preferably, the healthcare workers should engage their own communities or places where they are considered as peers, since studies have shown that interventions delivered by peers and community members were effective in improving outcomes (O’Mara-Eves et al., 2015). Engaging community members was successfully implemented by the CORE Group Polio Partners to improve routine immunisation coverage in Northern Nigeria (Usman et al., 2019). Finally, There is a need for the government and relevant stakeholders to strategize on how best to tackle COVID-19 related misconceptions in the North-Western region before the inevitable arrival of vaccines.

### Limitations and strengths

One major limitation for our study is sampling bias. Our study sample composed of respondents reached via WhatsApp, and thus, non-users of the internet, which form the majority of the population, are not part of the study population. The strength of the study is the large sample size. Therefore, the strong association between level of education and many dependent variables in the study could be used to set research questions for future studies. Furthermore, it is reasonable to design campaigns and policies targeting internet users (about 2.4 million (National Bureau of Statistics, 2019)), who mainly reside in the more urbanised areas of the state where COVID-19 is spread.

## Data Availability

The data that supports the findings of this survey are available from the corresponding author and may be provided upon reasonable request

## Supplementary material 1

### Tables

**Supplementary Table 1:** Answers to COVID-19 related knowledge questions by level of education

| Level of education | Answers % correct |  |  |
| --- | --- | --- | --- |
|  | Q2 | Q5 | Q6 |
| None/others | 36 | 18 | 27 |
| Junior secondary school and below | 77 | 31 | 40 |
| Senior secondary school | 52 | 50 | 54 |
| Diploma/NCE | 64 | 43 | 57 |
| HND/Bachelors degree | 65 | 42 | 69 |
| Master's/Professional degrees and above | 56 | 41 | 74 |

**Supplementary Table 2:** Practices of the respondents based on attitudes

| Variables | Practice (% yes) |  |  |
| --- | --- | --- | --- |
|  | Going to crowded places | Wearing of face mask | Washing hands |
| **Response to questions** |  |  |  |
| Q13. Final success in control (Yes) | 40.03 | 66.01 | 90.03 |
| Q13. Final success in control (No) | 41.02 | 56.41 | 71.79 |
| Q13. Final success in control (I don't know) | 36.62 | 59.15 | 84.50 |
| Q14. Confidence in government (Yes) | 36.72 | 67.58 | 91.21 |
| Q14. Confidence in government (No) | 47.14 | 58.10 | 81.90 |

**Supplementary Table 3.** Knowledge score of the respondents based on attitude and practices

| Variables | Knowledge score |
| --- | --- |
| **Response to questions** |  |
| Q13. Final success in control (Yes) | 9.69 ± 1.77 |
| Q13. Final success in control (No) | 9.76 ± 1.81 |
| Q13. Final success in control (I don't know) | 8.34 ± 2.93 |
| Q14. Confidence in government (Yes) | 9.73 ± 1.73 |
| Q14. Confidence in government (No) | 9.14 ± 2.37 |
| Q15. Going to crowded places (Yes) | 9.52 ± 2.04 |
| Q15. Going to crowded places (No) | 9.55 ± 1.97 |
| Q16. Wearing of face mask (Yes) | 9.82 ± 1.81 |
| Q16. Wearing of face mask (No) | 9.55 ± 1.97 |
| Q17. Washing hands (Yes) | 9.64 ± 1.86 |
| Q17. Washing hands (No) | 9.55 ± 1.97 |

**Supplementary Table 4.** Relationship between having a COVID-19 related misconception and COVID-19 related attitudes and practices

|  | No misconception | At least one |
| --- | --- | --- |
| Final success in control (Yes) | 92 | 83 |
| Confidence in government (Yes) | 80 | 69 |
| Going to crowded places (Yes) | 34 | 41 |
| Wearing of face mask (Yes) | 72 | 63 |
| Washing hands (Yes) | 93 | 88 |

## Supplementary material 2

**Hausa language version of the questionnaire used**

1. Wane Yankin shekaru ka/ki ke daga cikin wadannan?

a. 16-24
b. 25-39
c. 40-59
d. 60 abinda yayi sama
2. Menene jinsin ka?>

a. Namiji
b. Mace
3. Kana/kina da aure?

a. A’a
b. Eh
c. Na dai taba aure
4. Menene matakin ilmin ka/ki?.

a. Firamare
b. Karamar Sakandare
c. Babbar Sakandare
d. Diploma/NCE
e. HND/Digirin farko
f. Digiri na biyu
g. Digiri na musamman
h. Digirin digirgir (PhD)
i. Babu
j. Wani daban_____
5. Menene aikin ka/ki?.

a. Aikin karfi
b. Sanaár hannu ko kasuwanci
c. Ma’aikacin gwabnati (Mataki na 12 zuwa kasa)
d. Ma’aikacin gwabnati (Mataki na 13 ko sama
e. Na aje aiki
f. Dalibi
g. Matar aure
h. Bana aiki
i. Wani daban______
6. Karamar hukumar da ka/ki ke zama a halin yanzu _______
7. Alamomin da ake gane cutar COVID-19 sun hada da masassara, yawan gajiya da tari marar majina Gaskiya ne, Ba gaskia bane, Ban sani ba
8. Sabanin mura da aka saba, masu cutar COVID-19 ba su da cunkucewar hanci, zubar majina da atishawa Gaskiya ne, Ba gaskia bane, Ban sani ba
9. Yanzu cutar Korona ba ta da magani, sai dai gano cutar da wuri da shan magani mai taimakama jiki, zai iya taimaka ma mafi yawan masu cutar su warke. Gaskiya ne, Ba gaskia bane, Ban sani ba
10. Ba kowane mutum da ya kamu da cutar COVID-19 ne zai samu matsanancin rashin lafiya ba. Amma Tsofaffi, da masu ciwace ciwace da suka dade da masu matsananciyar kiba su suka fi shiga mawuyacin hali ta dalilin cutar Gaskiya ne, Ba gaskia bane, Ban sani ba
11. Cin naman dabbobin daji da muamala dasu zai iya kawo kamuwa da kwayar halittar COVID-19 Gaskiya ne, Ba gaskia bane, Ban sani ba
12. Mutanen da suke da kwayar halittar COVID-19 wadanda babu zazzabi tare dasu ba zasu iya sanya ma lafiyyayun mutane ba cutar ba Gaskiya ne, Ba gaskiya bane, Ban sani ba
13. Cutar COVID-19 tana yaduwa ne ta hanyar feshi ta baki ko ta hancin wadanda suke dauke da kwayar halittar Gaskiya ne, Ba gaskiya bane, Ban sani ba
14. Game garin al’umma zasu iya sanya takunkumin fuska don su kare kansu daga kamuwa da cutar COVID-19 Gaskiya ne, Ba gaskiya bane, Ban sani ba
15. Bai zama dole ga yara kanana da matasa su yi kokarin kare kansu daga cutar ta COVID-19 ba Gaskiya ne, Ba gaskiya bane, Ban sani ba
16. Don kariya ga kamuwa da cutar COVID-19, dole mutum ya kiyayi zuwa wurare masu cunkoso kamar kasuwanni kuma ya kiyaye shiga ababen sufuri na haya Gaskiya ne, Ba gaskiya bane, Ban sani ba
17. Killace wadanda suka kamu da cutar COVID-19 tare da yi masu magani, hanya ce da za a iya rage yaduwar cutar Gaskiya ne, Ba gaskiya bane, Ban sani ba
18. Mutanen da sukayi mu’amala da masu cutar COVID-19 dole ne a killace su da gaggawa a wurin da aka tanada. Lokacin da ake kula da wadannan mutanen mafi yawanci kwana 14 ne Gaskiya ne, Ba gaskiya bane, Ban sani ba
19. Ka/kin yarda cewa za a iya maganin cutar COVID-19 cikin nasara daga karshe? Na yarda, ban yarda ba, ban sani ba
20. Ka/ki na da yakini cewa Nigeria za ta ci nasarar yaki da cutar COVID-19? Eh, a’a
21. Cikin sati biyu da suka wuce, shin ka/kin je wuri mai cunkoso? Eh, a’a
22. Cikin yan kwanakin da suka wuce, shin ka sanya takunkumin fuska kafin kabar gida? Eh. A’a
23. Cikin yan kwanakin da suka wuce, kana wanke hannunka da ruwa da sabulu ko sinadarin kasha cututtuka na hannu a kai a kai? Eh, a’a
24. Kana ganin cewa kwayar halittar COVID-19 an kirkira ce ta wata gwamnati ko kungiyar yan taadda? Eh, a’a, ban sani ba
25. Kana gani cutar COVID-19 ta samu ne dalilin cigaba na kimiyya irin su 5G? Eh, a’a, ban sani ba
26. Kana ganin cewa cutar COVID-19 anyo ta ne domin a rage yawan mutanen duniya? Eh, a’a, ban sani ba
27. Wace hanya ce mafi rinjaye da ka/ki ke samun bayanai game da cutar COVID-19Ta wace hanya kake samun bayanai game da cutar COVID-19? Zabi duka kafofin da ka ji labarin cutar

a. Tashar FM
b. Tashar radiyo da ba FM ba
c. Tashar talabijin (KTTV ko NTA)
d. Tashar talabijin na satilayit na kasa (National, Channels, TVC etc)
e. Tashar talabijin na satilayit na kasashen waje
f. WhatsApp
g. Ta hanyoyin sada zumunta wadanda ba whatapp ba
h. Ta yanar gizo
i. Ta hanyar ma’aikatan lafiya
j. Ta hanyar yan’uwa
k. Abokai
l. Ma’aikatan lafiya na al’umma
m. Masu wayar da kan jama’a
n. Shuwagabannin al’umma
o. Malaman addini
p. Masu maganin gargajiya
q. Ungo zoma
r. Al’ummar gari
s. Yi bayani in ba hanyar a sama_____
28. Wace hanya ka fi yadda da ita domin samun labaran cutar korona? (Zabi daya ko fiye da haka)

a. Tashar FM
b. Tashar radiyo da ba FM ba
c. Tashar talabijin (KTTV ko NTA)
d. Tashar talabijin na satilayit na kasa (National, Channels, TVC etc)
e. Tashar talabijin na satilayit na kasashen waje
f. WhatsApp
g. Ta hanyoyin sada zumunta wadanda ba whatapp ba
h. Ta yanar gizo
i. Ta hanyar ma’aikatan lafiya
j. Ta hanyar yan’uwa
k. Abokai
l. Ma’aikatan lafiya na al’umma
m. Masu wayar da kan jama’a
n. Shuwagabannin al’umma
o. Malaman addini
p. Masu maganin gargajiya
q. Ungo zoma
r. Al’ummar gari
s. Yi bayani in ba hanyar a sama_____

